# Strong impact of closing schools, closing bars and wearing masks during the COVID-19 pandemic: results from a simple and revealing analysis

**DOI:** 10.1101/2020.09.26.20202457

**Authors:** Polly Matzinger, Jeff Skinner

## Abstract

Many complex mathematical and epidemiological methods have been used to model the Covid-19 pandemic. We took a different approach. Making no assumptions, we simply plotted cases, hospitalizations and deaths, on a log_2_ Y axis and a linear date-based X axis, and analyzed them using segmented regression. The data fit straight lines with correlation coefficients ranging from 92% - 99%, and these lines broke at interesting intervals, revealing that school closings dropped infection rates in half, lockdowns dropped the rates 3 to 4 fold, and other actions (such as closing bars and mandating masks) brought the rates even further down. Hospitalizations and deaths lagged, but paralleled cases. The graphs, which are easy to read, have several implications for modeling and policy development during this and future pandemics. Overall three interventions had the most impact: closing schools, closing bars and wearing masks: a message easily understood by the public.

**One Sentence Summary:** Regression analysis showed that closing schools, closing bars, and wearing masks had major effects on infections, hospitalizations and death rates in the US during the Covid-19 pandemic.

## Main Text

As of August 1^st^, 2020, the Covid-19 pandemic has been in effect for about six months in the US. Infection rates have climbed, fallen and climbed again, as different state governments ordered interventions to fight the pandemic, rescinded them to open their economies, and then (in some cases) reinstituted them as infections resurged. In this study, we analyzed the effects of those interventions using a simple approach. We started by asking if a pandemic in its earliest stages, when the population is completely naïve, might be described by simple kinetics similar to those that model chemical reactions in their earliest stages, before the increasing concentrations of reaction products begin to impinge on the reaction rate. Making no assumptions, we simply plotted the data for Covid-19 cases in several states in the USA as the pandemic evolved. We confined our analysis to the USA for several reasons. First, we did not think it wise to compare countries with different types of health systems. Second, in the absence of a coordinated federal response, different states made different policy decisions, on different dates; allowing for detailed local comparisons of states with strict and early lockdowns, states with no restrictions at all, and states with similar policies implemented at different times. Because of these state-to-state differences, we thought that an analysis done at the granular state level might be able to distinguish the results of different interventions from one another, and show which were most effective.

Our earliest plots indicated that the daily increases in cases during the initial days of the pandemic follow a purely exponential trajectory, regardless of country, state or county. We therefore plotted the data on a log_2_ Y axis, choosing log_2_, rather than the more standard log_10_, because it makes the doubling time easy to calculate (e.g. if a plot of new cases per day is a straight line and passes through three horizontal lines [2^3^] in a period of 15 days, then the doubling time is 5 days. We also saw that the curves could be rendered as a series of straight lines that visually seemed to “break” into new straight lines with new slopes that seemed to correlate with changing government policies. To add rigor to our visual results, we used segmented regression analysis, a statistical method for determining discontinuous changes in slope. We found, for the first few months of the pandemic, that the data fit straight lines with R values usually greater than 92% - 99%, and that the breaks (slope changes) occurred at interesting and revealing intervals. For example, school closings reduced the rate of infections by half or more, with a 6-14 day delay. Lockdowns reduced the rates of infections by 2 to 4 fold, with a similar delay. Further actions (such as mandating masks or closing bars) similarly brought the rates further down with somewhat more variable lag times.

Deaths, as expected, were a lagging indicator, but closely paralleled the other two main parameters (cases and hospitalizations) about one week later. This method remained useful even in later stages of the pandemic, as the country began to open up, as it enabled visualization of the effects of relaxing restrictions, and showed the mitigating effects of late interventions. The graphs, which are easy to read, show how various individual policy decisions reduced or increased the rates of new Covid cases, hospitalizations and deaths; and can potentially serve as the groundwork for future policy decisions during this and future pandemics.

## Results

### Outlining the process for one state

We begin here with data from a single state, Maryland (MD), the home of the NIH, where we assumed we would have access to precise data (an issue with some states). We graphed cases, hospitalizations and deaths on a log_2_ Y axis, and applied segmented regression analysis to evaluate the accuracy of the visually estimated slopes and breakpoints.

Figure 1 displays a progression of graphs for the state from March 6, 2020, when the first three cases were reported, to April 15^th^. This simple method appeared to be adequate to model the data during this initial stage of the pandemic, and revealed some features that were not obvious from other, more commonly used plots, most of which are plotted on linear Y axes^1-2^ or, at best, on axes of Log_10_ (ref 3).

**Figure 1.**
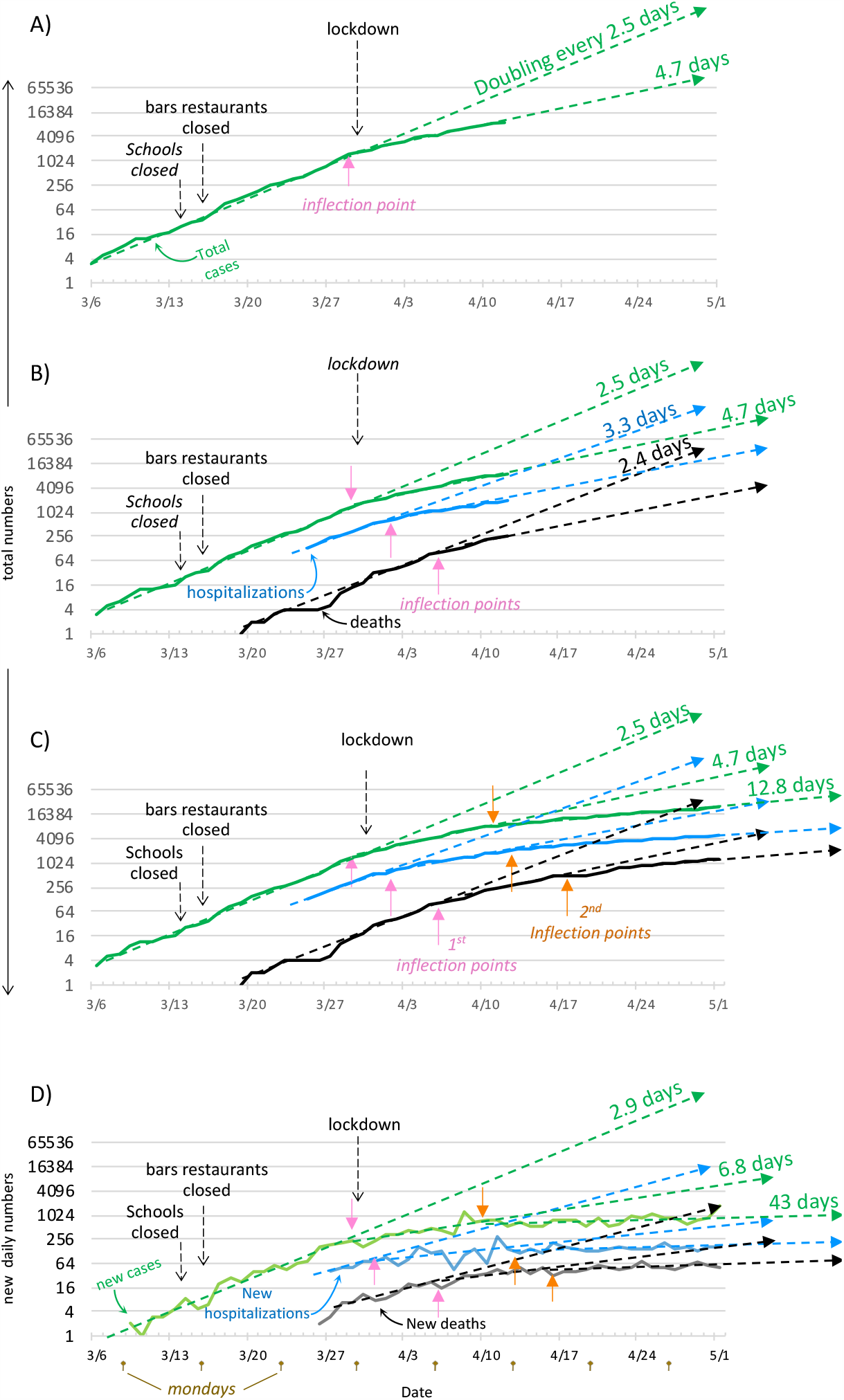
Laying out the process for one state (Maryland). **(A-C)** Graphs for total accumulating cases, hospitalizations and deaths. **(D)** graph of new daily cases, hospitalizations and deaths. Pink and Orange arrows =1^st^ and 2^nd^ inflection points respectively. **First rate drop:** Initially Covid-19 cases double every 2.5 days. Then, 15 days after schools, bars & restaurants are closed (and one day before lockdown), the doubling rate slows to every 4.7 days. **B) Other parameters also drop**. Hospitalization and death rates drop, 3 and 8 days respectively after the decline in cases. **C) 2**^**nd**^ **rate drop:** the lockdown is followed by a second rate decline to a doubling time of 12.8 days, for cases, hospitalizations and deaths at 12, 14 and 18 days later respectively **D) New daily cases:** new cases, new hospitalizations and new deaths follow similar patterns. The data are less smooth, partly due to the smaller numbers and partly because reporting is lower on Sundays and Mondays (brown arrows) and rebound later in the week.

Because the total number of Covid-19 cases (Figure 1A, solid green line) tended to increase exponentially, resulting in a fairly straight line on the log_2_ scale, we were able to determine from the slope (dashed green line) that the initial doubling time was 2.5 days. This rapid rate lasted until March 29, at which time it slowed to a doubling time of 4.7 days. This change occurred 15 days after schools closed, 12 days after bars and restaurants closed, and one day before the state lockdown, suggesting that the effect of one or both of the first two policy decisions was to reduce the doubling rate of cases to slightly more half of the original rate. Figure 1B shows that the slopes for hospitalizations and deaths followed the same pattern, with lag times of about three and eight days longer respectively than the lag times of positive cases. Figure 1C shows the effect of the lockdown in MD, which was followed twelve days later by an additional threefold change in doubling time, from 4.7 to 12.8 days, and similar rate reductions in hospitalizations and deaths with additional lags of 2 and 6 days respectively. Segmented regression analysis showed that the combined broken lines fit the data remarkably well, with R values well over 99 for total cases (99.89), for total hospitalizations (99.85) and total deaths (99.42). Figure 1D shows the data for daily new cases, whose slopes tend to change within a day of those for total cases, and undergo greater changes.

The fact that the inflection points for daily new cases often occur one day before those for total cases, rather than on the same day, and that the correlation coefficient value (96.99) is slightly lower than that for total cases, is likely due to the fact that daily new cases have a periodic dip on Sundays and Mondays, and then rebound slightly as the reporting catches up during the week. This variability results in a greater “fuzziness” that creates an uncertainty in the break points and a somewhat lower accuracy in the slopes. Nevertheless, the high correlation coefficients suggest that these slopes are good representations of the trends in the data.

### Effects of School Closings and of Lockdowns

Because Maryland closed bars and restaurants just three days after closing schools, it was 10 impossible to separate the effects of those two actions from each other. A recent study on closing schools^4^ ran into the same problem. In fact, the near simultaneous execution of several different interventions by many states and countries has made it difficult to assess the value of any one particular intervention. We therefore asked if any state had witnessed changes in slopes after closing schools but before taking any other major action, such as closing bars and restaurants, closing essential businesses, or giving “stay at home” orders. We found three states with no mask mandates (Georgia, Tennessee, Mississippi), that had spaced their other interventions far enough apart to enable us to examine the effects of each by itself. (Fig2)

Georgia (Figure 2A) officially closed schools on March 16^9^. However, the 16^th^ was a Monday, and we therefore chose to specify Saturday, March 14^th^, as the school closing date, as this was the first day that children did not attend school. Seven days later, on March 21^st^, Georgia’s initial doubling rate of 2.2 days for total positive cases dropped to a doubling time of 3.5 days. Two days after that, Georgia closed bars and restaurants, and this was followed eight days later by a second change to a doubling time of 7.6 days. Two days after this second rate drop, the governor ordered a lockdown^11^, which was followed by a third decline in the slopes after a lag of nine days. The observation that each change in doubling times occurred before the next government intervention suggests that each intervention is followed by its own rate change and that additional interventions have additional effects.

**Figure 2:**
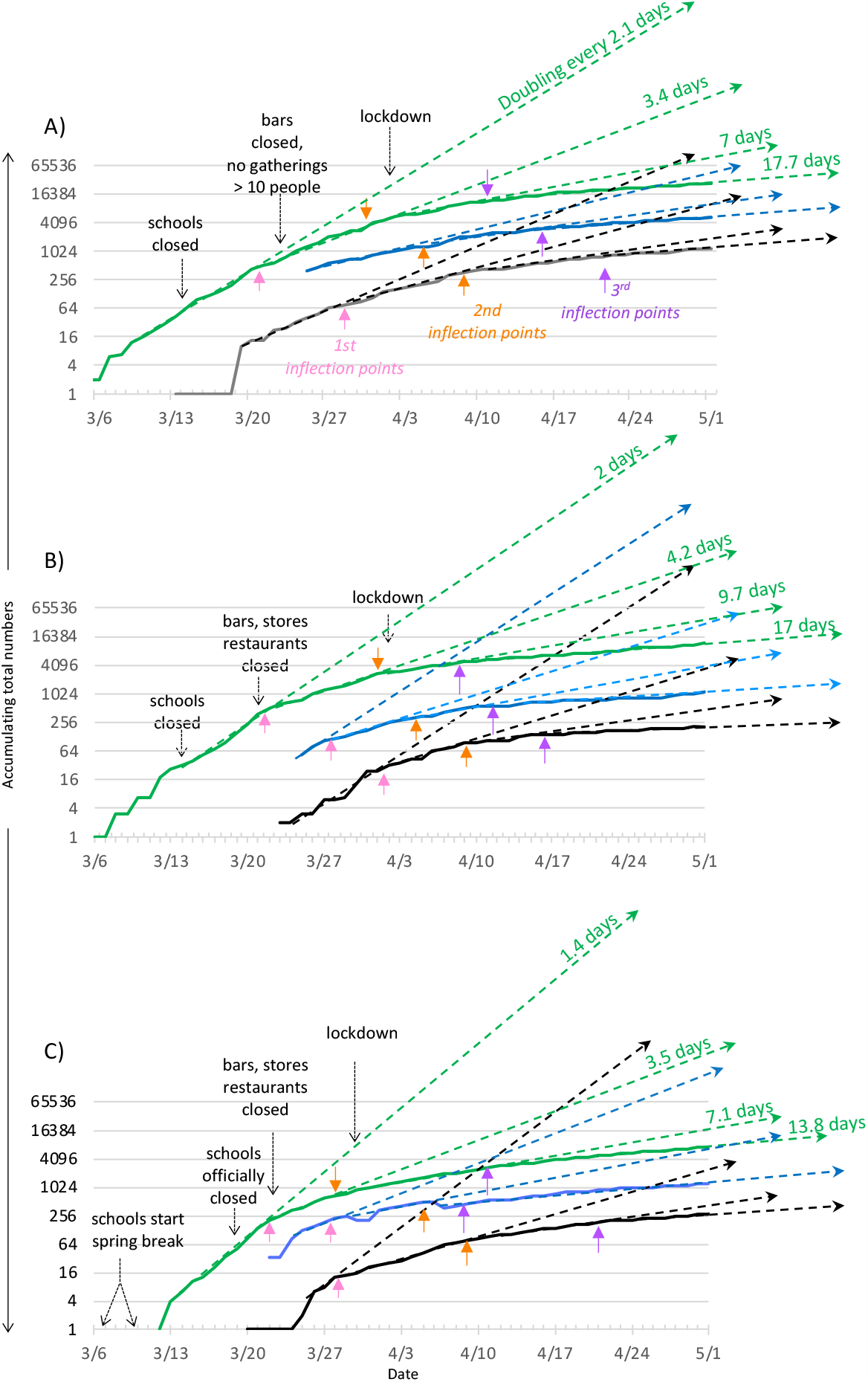
the effect of closing schools: graphs cover three states from 3/6/2020 to 5/1/2020. Colored arrows point to inflection points. Pink = 1^st^, orange = 2^nd^ and purple = 3^rd^ inflection point. A) **Georgia:** Total cases and death rates decline 8 and 14 days, respectively, after school closings. Cases, hospitalizations and deaths drop again 11, 14 and 17 days, respectively, after closing bars, and again, with similar lag times, after lockdown. **B) Tennessee:** Total cases decline 8 days after school closings, again 11 days after non- essential businesses close, and again 7 days after lockdown. Hospitalizations and deaths follow with typical lag times. **C) Mississippi:** Case rates drop about 2 days after official school closings (10-12 days after the start of spring break), again 10 days after non-essential businesses close, and again 14 days after the lockdown. Hospitalizations and deaths follow with variable lag times.

Two days after this second rate drop, the governor ordered a lockdown^11^, which was followed by a third decline in the slopes, after a lag of nine days, to a doubling time of 17.7 days. The observation that each change in doubling times occurred before the next government intervention suggests that each intervention is followed by its own rate change and that additional interventions have additional effects.

Tennessee (Figure 2B) also closed schools on March 14^th^ and saw a twofold drop 8 days later, the same day that bars, restaurants and non-essential business were closed. This second intervention was followed eleven days later by another twofold drop in rate. One day after this second drop, the governor issued a “stay at home” order^14^, which correlated with a third drop 7 days later.

Mississippi (Figure 2C) was a puzzle at first, because the first rate drop occurred only two days after the official order to close schools^5^. Rather than dismiss this as a simple outlier - it was the only state among the 50 that had such a short lag time - we delved into other possible explanations, thinking that perhaps we were simply wrong, and some aspect of human behavior other than school closings was responsible for the first sets of rate reductions. Perhaps, Americans had simply started going out less, as suggested by the phone mobility data from Apple, Google and Cuebiq^6-8^.

However, an analysis of the mobility data for Mississippi in comparison with other states (Supplemental Figure 1), showed no change in mobility that could underlie Mississippi’s early rate change. We therefore looked for other factors, and discovered that spring breaks for Mississippi colleges^9^ and K-12 schools^10^ had almost all begun earlier than other states, with slightly variable start dates from March 6 to March 10. Although the K-12 breaks normally last only one week, the governor asked schools to extend their spring breaks because of the virus^11^, and nearly all of the schools did not go back into session before he officially closed them on the 18^th^ of March^5,12^. Consequently, the drop in the rate of cases that occurred on March 20^th^ most likely resulted from the fact that children had stopped attending school 10-14 days earlier, rather than from the March 20^th^ executive order to close schools.

Thus, for states that separated their government actions by a week or more, we were able to assess the effects of those actions individually. Closing schools appeared to cut the rate of infections in half, beginning 8-14 days post-closure, and similarly reduced hospitalizations, but with lag times 2-4 days longer than for positive infections. Deaths followed suit, lagging an additional 7-11 days.

Closing bars and restaurants and ordering lockdowns similarly reduced the rates of infections, with similar lag times. Cumulatively, these interventions resulted in 7-10 fold drops in the rates of infections, hospitalizations and deaths. Nevertheless, even when combined, these effects were frequently insufficient to result in a steady decrease in the daily tallies of new infections. For that we needed masks.

## Mask Mandates

Although the federal government never mandated masks, and many government officials continued to eschew them, a few state governors wrote orders for their constituents to wear masks. Unlike school closings, which occurred all over the US in the space of about two weeks, from March 6 (Mississippi) to March 23^rd^ (Idaho), mask mandates were put in place over a longer time period, from March to July. This gave us an opportunity to evaluate both early and late mandates. We found that they were about equally effective (Figures 3 and 4).

**Figure 3:**
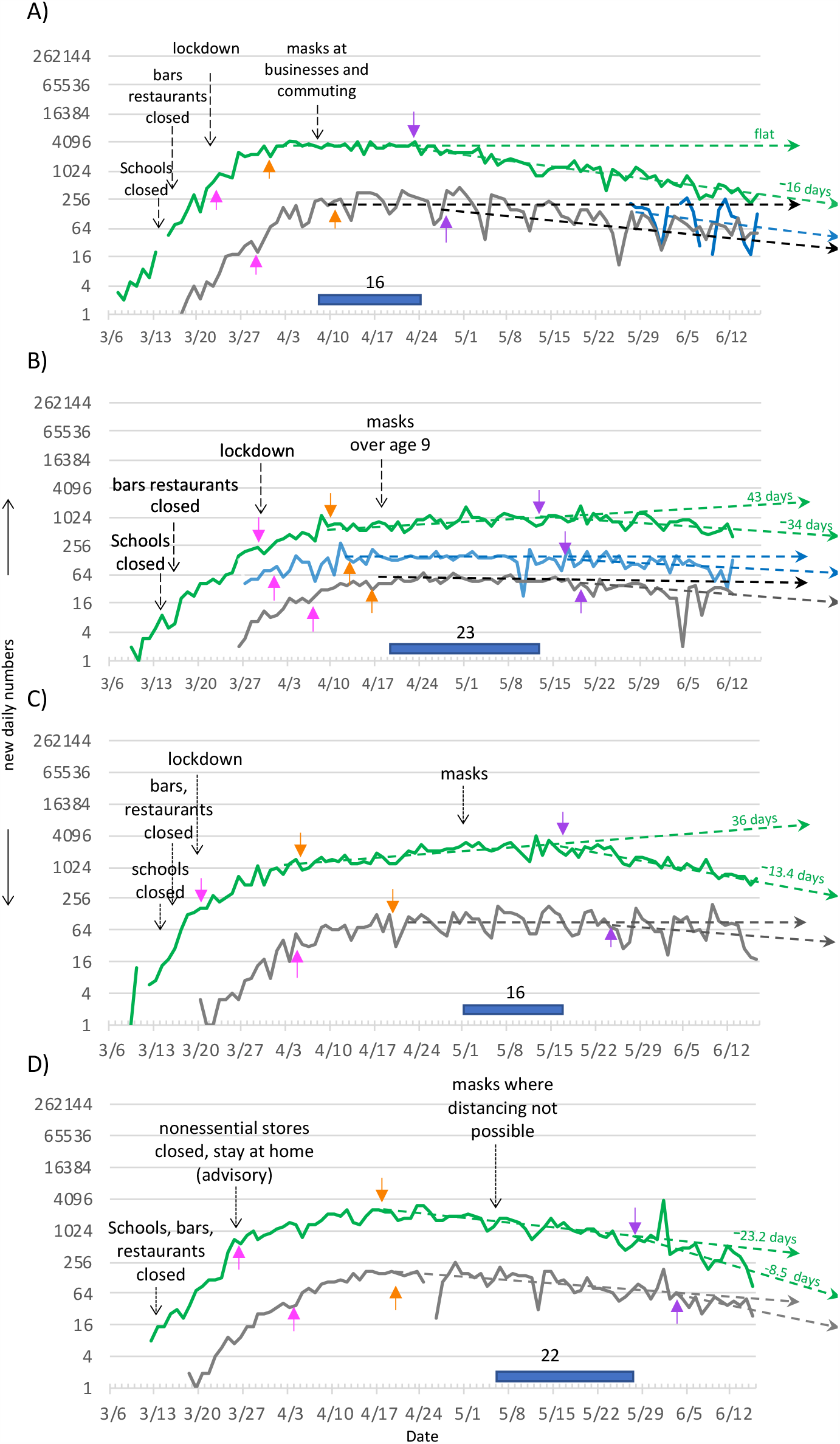
the effect of wearing masks: graphs depict new daily numbers for four states that mandated masks at different times from April 8 to May 6, 2020. Pink, orange and purple arrows point respectively to 1^st^, 2^nd^, 3^rd^ inflections points. **A) New Jersey:** 16 days after mask mandate on 4/8 the slope drops about 2 fold. After this point, daily new cases drop by half every 16 days. **B) Maryland:** 23 days after mask mandate, on 4/18 the slope drops 1.5 fold to a halving rate of 34 days. **C) Illinois:** 18 days after mask mandate on 5/1 the **s**lope declines about 2 fold, to a halving rate of 13.4 days. **D) Massachusetts:**18 days after mask mandate on 5/6, an already declining slope drops about 2 fold further, to a halving rate of 8.5 days.

**Figure 4:**
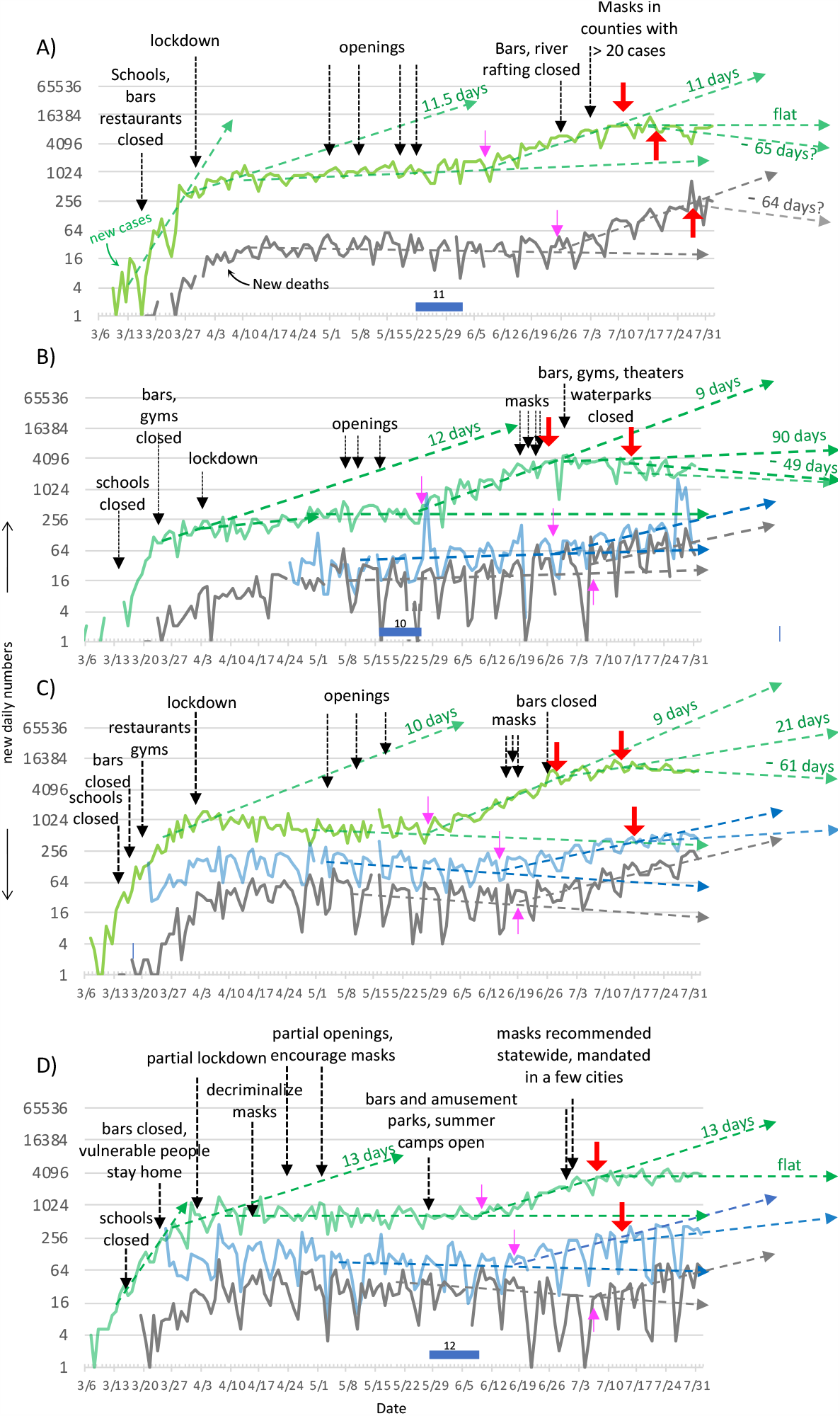
infection rates after opening mimic post- school-closing rates, and are flattened by wearing masks and closing bars. Small pink and large red arrows point respectively to beginnings of surges and reductions in infections. **A) Texas:** May1: opening almost everything (but not bars). May 8: personal care services open. May 18: gyms open. May 22: bars. On 6/16 Texas suddenly added 14 open. **B) Arizona openings**: May 8: many businesses open. May 11: restaurants open. May 16: everything else opens (including bars). **C) Florida openings**: May1: some counties open. May 11: Palm beach opens. May 18: Miami- Dade and Broward counties open. **D) Georgia openings**: April 24: personal services, gyms open. May 1: opening just about everything except bars and amusement parks. May 28: Bars, amusement parks, overnight camping open.

Figure 3 shows the effects of early mask mandates from four states that ordered the use of masks at times spanning a month, from April 8^th^ to May 6^th^. We depict new daily numbers, as total numbers were high enough by this time to be slow to show intervention effects. Each state’s mandate was followed by a drop in the rates of infections, and then by drops in the rates of hospitalizations (where reported) and deaths, showing that the effect of mandating masks is to drop the slopes about 2 fold. As the mandates stepped through time, the rate reductions followed in concert, with later mandates correlating with later inflection points. This supports the view that these rate reductions were due to wearing masks, rather than other potential changes in mobility^6-8^ or behavior.

The lag times for the effect of masks (16-23 days, as indicated by dark blue bars near the X axis) seemed somewhat longer than those that followed closing schools, bars and restaurants, or ordering lockdowns, perhaps because it takes time for people to obtain masks and get into the habit of wearing them, whereas a school or a business can be closed in a day.

An exception to the effectiveness of mask wearing was seen in New York, which implemented a mask mandate during its lockdown, when new daily cases were already being cut in half every

## Openings

As states reopened their economies, and experienced surges in new infections, we found that the new infection rates showed the effects of previous interventions that remained operational, as well as clear effects of interventions that were implemented later. For example, figure 4 shows data for four states (Texas, Arizona, Florida and Georgia) that opened in the middle of May. We chose these four because they had not previously mandated masks, in order to determine the effect of late mask mandates. These states had flattened or even reversed their slopes by closing schools, bars and restaurants, and non-essential businesses. All but Georgia had also implemented strict stay-at-home orders (Figure 4A - C)

The first effect we observed was the lasting impact of school closings. As infection rates surged following reopening of state economies, the benefit from having closed the schools persisted. In all four states, as schools remained shuttered for the summer, the rates of new infections never recouped the initial doubling times of 2-3 days, instead returning to the slower doubling times of 9-11 days that had followed the school closings, despite the fact that all other restrictions had been lifted. This suggests that the effect of each intervention endures as long as that intervention remains place, regardless of other actions that may be implemented or rescinded.

The second thing we noticed was that the post-opening surges seemed to be strongly correlated with the opening of bars. Regardless of the timing or sequence of other relaxations, opening bars was followed 11-12 days later by surging infection rates. Texas, for example (Figure 4A), opened many businesses, including restaurants from May 1^st^ to May 18^th^, with little repercussions on infection rates. On May 22^nd^, bars were opened (along with aquariums, zoos, rodeos, bowling alleys, bingo halls and skating rinks) and 11 days later, infection rates surged.

Arizona (Figure 4B) opened everything over an 8 day period, with bars opening only on the last day (May 18^th^). As with Texas, Arizona’s surge of new cases also began 11 days later. Georgia (Figure 4D) carefully opened its economy over a period of more than a month, with no ill effects until it opened bars on the last day, along with amusement parks and overnight camping. It saw a surge starting 12 days later. In Florida each county decided individually when to open, making it impossible to determine the effect of opening bars in the Sunshine State, but for the other three states, the lag from opening bars to surges in infection rates was remarkably consistent (11-12 days), with only 1 day’s difference among the three.

Thirdly, the data suggested that the simple combination of closing bars and wearing masks was a highly effective way to clamp down on surging infection rates. Each of these measures brought the infection rates down by twofold, for a cumulative reduction of fourfold in new infection rates. States that implemented both interventions, put surging infection rates into decline. Texas, Arizona and Florida (Figure 4 A-C) all carried out both interventions, though in different orders. In all three states, surging doubling times of 9-11 days sank to halving times ranging from 49 (Arizona) to 65 (Texas) days.

In contrast to the other three surging states, Georgia’s governor neither closed bars nor mandated masks (Figure 4 D). However, he did encourage mask wearing, and several mayors mandated them for their cities, covering about 20% of the state’s population. This mask mandate was followed about ten days later by a twofold reduction in the rate of new cases. Nonetheless, new cases did not go into decline, but instead flatlined. The major difference between Georgia and the three states that dropped their post-opening surge rates to declining ones is that the more successful states not only mandated mask wearing, they also closed their bars.

Together the data from these four states suggest that bringing cases down to manageable levels might not require shutting down an entire economy. The combination of closing schools, closing bars, and wearing masks may be enough.

## Discussion

As countries and states debate the best way to prevent Covid-19 infections, and as they contemplate re-opening schools in the fall, the analysis of the information thus far gleaned about this new pandemic is vital to those decisions. There have been several previous attempts to correlate various government interventions with rates of SARS-Cov-2 infections, and predict which might be most effective^13-18^. These studies, however, have mostly taken a global view, and this is not necessarily the most productive for the US. Although a global view has the advantage of large numbers, it has the disadvantage of combining disparate interventions, done at different times in different places, into one global soup, in an attempt to find common themes.

We honed down instead to the granular data at the state - and sometimes city - level, painstakingly combing governors’ executive orders and press releases (and those of mayors when necessary) to find examples of states that had implemented each of their interventions sufficiently far apart to enable us to distinguish the effect(s) of each individual intervention.

A second difference between this and previous studies is the method we used. A maxim among scientists is to use the simplest method that explains the data. Our decision to plot the data, as they came, without any assumptions, on a Log_2_ Y axis, allowed us to see the straight lines in the data, find the slopes of those lines, and the inflection or “break” points where those slopes changed. The human eye, coupled with human intuition is a powerful tool, and can guide a simple mathematical approach. We therefore used segmented regression analysis to test our visually-based hypotheses about breakpoints, slopes and doubling times. The high R values for the segmented slopes highlight the accuracy of the visually based breakpoints, despite the variability of the data of new daily cases known to occur because of the “weekend effect” of low reporting, and of low initial numbers.

The results, demonstrate that, other than full lockdowns, three government interventions had the most impact on the rates of Covid-19 infections: closing schools, closing bars and wearing masks.

### Schools

Closing schools not only flattened the early curves of rapidly rising numbers of infections in March (Figure 2), it prevented a return to those initial rates after states opened up their economies in mid-summer - a time when schools remained closed (figure 4). This demonstrates that the effect of a particular intervention lasts as long as that intervention is in place, regardless of other interventions that might be mandated and/or rescinded.

Although it could be argued that the mid-summer surging rates of infections were lower than the initial rates in March because of increased resistance in the US population, this is unlikely. At the time (early to mid-June), the number of confirmed cases in the US – just under 2 million - constituted a mere 0.6 percent of the population^19^. Even if the true infection rate were 8-10 times higher, as underscored by a recent study^20^, the increase to 6% would still be too small to account for the reduced rates of post-opening infections, which closely mimicked the early post-school-closing rates.

These findings help to resolve the conflicting conclusions of two previous reports. In contrast to two early predictive models suggesting that closing schools would bring down Covid-19 infection rates, a study covering the effects of government interventions in six countries^21^ concluded that closing schools had no effect in the US. However, the authors analyzed each of those countries as a whole, which is not useful in the case of the US, which had no national policy on the matter. The second study^22^ analyzed individual states, and concluded that closing schools had a major impact. However, in evaluating all the states together, the authors noted that they were unable to discount the potential effects of other interventions. Our study, like theirs, shows that closing schools had a major effect, and supports this idea with the results from the small set of states (Figure 3), where the effect could not be explained by other interventions.

### Bars

The effect of closing and opening bars became evident in those states that opened their economies in stages (Figure 4). Although most states closed bars and restaurants simultaneously during their early shutdowns, some opened them at different times during the re-openings. We found that, regardless of other relaxations, new infections surged beginning 11-12 days after bars were opened, and fell once again about 8 days after bars were re-shuttered. This suggests that closing (and re-opening) settings that might not be conducive to social distancing has more impact on new infection rates than would opening other types of businesses (dog groomers, markets, hardware stores; even restaurants).

### Masks

Akin to school closings, the beneficial effect of wearing masks was evident during the early period when states were closing down (Figure 3), and also later during post-opening surges. Under both conditions, those states that mandated masks^23^ saw about a twofold reduction in the rate of new cases. Although political controversy reduced the proportion of the population that wears masks, those states that combined masks with bar closures saw their post-opening surge rates drop to declining ones (Figure 4). We surmise that better adherence to the mask mandates would likely have generated even larger decreases in infection rates.

One exception to the effect of early mask mandates was evident in the data from New York (Supplemental figure 2), which mandated masks after it had already turned its rapidly rising infection numbers into declining ones. The mask mandate had no further suppressive effect on infections. However, when New York later opened its economy, and infections began to rise, that increase was remarkably lower than those of states that opened without mask mandates in place (figure 3), suggesting that the early mask mandate had a long lasting effect.

Several studies suggested that early interventions have more effect than later ones. However, our data show that late interventions can be just as effective as early ones. Compare, for example, the twofold reductions in infection rates that follow early mask mandates in Figure 3, with the similar reductions seen in figure 4 - with mandates made months later. Although the numbers of cases at the time of a late intervention may be larger, setting a higher baseline, the proportional rate reductions are about the same. Whether flattening a curve at 10 or1000 new cases per day, these interventions have a major effect that can lead, with time, to lowering infections to the same level.

Finally, Figures 1-4 show that rates of new daily hospitalizations and new deaths seem to parallel those of new cases, with lag times of about three days to a week for hospitalizations, and a further lag of 3 - 10 days for deaths. Although improvements in medical care may start to change this picture^24-26^, it currently appears that a fairly constant proportion of infected people will become ill and die. To counter this, we need a global set of effective policies to reduce new infections, as reductions in hospitalizations and deaths will follow. In the absence of a uniform US federal policy, we have been able to use the different state policies to determine the value of different interventions. Our analysis shows that, when combined, the three most powerful interventions - closing schools, closing bars and wearing masks - successfully flattened initial infection rates, and turned post-opening surges into declining ones, suggesting that the country could perhaps open up its economy safely if it kept bars and schools closed, and required the strict wearing of masks in public

### Coda

If, in a future pandemic with a new infectious agent, governing bodies decide to base decisions on analyses of rates of infections, hospitalizations and deaths, we will need to eliminate the weekend effect, which results in underreporting on weekends and Mondays, and over-reporting later in the week, in every state examined. This can delay the determination of a rate change for a week or more, because many data points must accumulate to reliably assign a breakpoint and new slope. As there is already a biological delay from infection to the onset of symptoms, and a further delay before testing, any additional delay prevents the timely implementation of needed interventions. In a rapidly evolving pandemic, reliable data are crucial to the outcome, and every day counts.

## Data Availability

all data and computation programs are freely available on request

## Acknowledgements

We thank Tim D. Blood of the Maryland Department of Health for pulling and cleaning data, Gabrielle Bains and Yvonne Rosenberg for scientific and editing suggestions, the science writer, David C. Holzmann for improving flow and clarity, Mihalis Lionakis and Robert Munford for critical review, and Steve Holland for support, insightful questions and occasional butt kicks.

## Funding

this work was entirely funded by the Division of Internal Research, National Institute of Infectious Diseases, NIH.

## Author contributions

Polly Matzinger conceived the graphing model, researched all the state interventions, wrote the paper and created the figures. Jeff Skinner wrote the programs to compute the statistical analysis and wrote the statistical analysis methods section.

## Competing interests

Authors declare no competing interests.

## Data and materials availability

All data are available in the main text and the supplementary materials. Code used for segmented regression analysis freely available on request.

## Supplementary Materials

### Materials and Methods

#### Data acquisition

Data for different US states were taken from those published online by the individual State Health Departments, from “the Covid Tracking Project”^27^ at the Atlantic under a <u>Creative Commons CC BY-NC-4.0 license</u>, from Statista^28^ and from the IHME^29^. For school closings, mobility data, and other government actions, we also relied on GitHub^30^ and then verified the dates with the various governors’ orders. If the date of official school closing was a Monday, we used the Saturday before as the first date that students were not in school. For Mississippi, we found that Colleges and Universities, and most K-12 districts had gone on spring break about a week and a half before the official Governor’s order to close the schools. We contacted the Mississippi department of Education, and most of the individual school districts, to find which, if any, had gone back to school in the interim between spring break and the official closing order, covering 97.2% of students in the state, and discovered that >87% of those students had not gone back to school, mostly because of local decisions by their mayors, school boards or boards of health. We consequently used the first day of spring break as the date of school closing in Mississippi.

For deaths, we report the combined “confirmed plus probable” when available.

#### Data analysis

Data were analyzed in two ways. First by visualization on a plot using a Log2 Y axis to find the approximate dates of breakpoints in the lines. Second, using segmented linear regression to find and estimate the breakpoints, and to compute regression slopes and confidence intervals. To this end, trends in the number of new cases, total cases and total deaths were estimated by segmented regression using the “segmented” package library in R following its authors instructions^31^. The independent variable (X) was always “number of days” starting from March 3^rd^ as Day 1. March 3^rd^ was chosen because none of the states examined had any COVID-19 cases, hospitalizations or deaths reported prior to March 3^rd^. Initial values for either 2 or 3 segment breakpoints were provided using the 33^rd^, 50^th^, and 66^th^ percentiles of date numbers spanning the range of the total deaths reported from that particular state. Total deaths were chosen because they typically had the smallest range of dates, so initial values chosen from the range of total deaths would usually work for the positive cases and the hospitalizations. If only 2 breakpoints were needed, then only 33^rd^ and 66^th^ percentiles were used. In rare cases where these initial values would not work to fit a particular variable (e.g. hospitalization or new cases), initial values would be chosen manually based on the date range of that variable. Estimated breakpoints were generally robust to the initial values chosen, and would typically yield the same estimated breakpoints even when substantially different initial values were provided. Estimated breakpoints were reported in number of days with their standard errors and their predicted date. Slopes for the resulting 3 or 4 regression segments were also reported with their adjusted 95% confidence intervals and their doubling times, calculated as 1/slope. The 95% confidence intervals can be compared against zero, to determine if slopes are increasing or decreasing, and they can be compared against each other to determine if one slope has significantly flattened or significantly increased relative to the previous slope.

#### Visualizations

Graphs follow the principles of Edward Tufte^32^ as closely as possible.

## Supplemental data

**Supplemental Figure 1.**
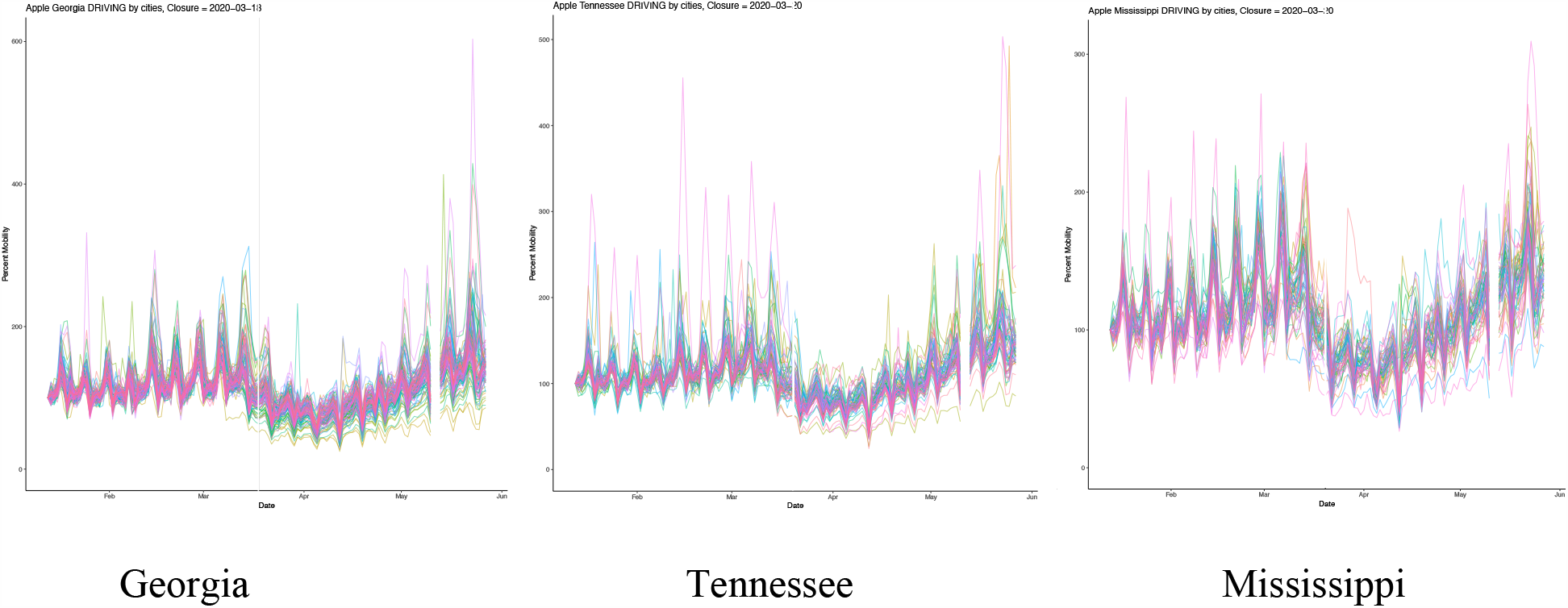
Apple Mobility data for Georgia, Tennessee and Mississippi: Graphs show how much cell phones moved by driving each day. Different colors represent different counties in each state. Overall, phones in all 50 states started moving less in mid march. (the small spike in mid February is Valentine’s day weekend). There is no obvious difference im the mobility data for the three states.

**Supplemental Figure 2.**
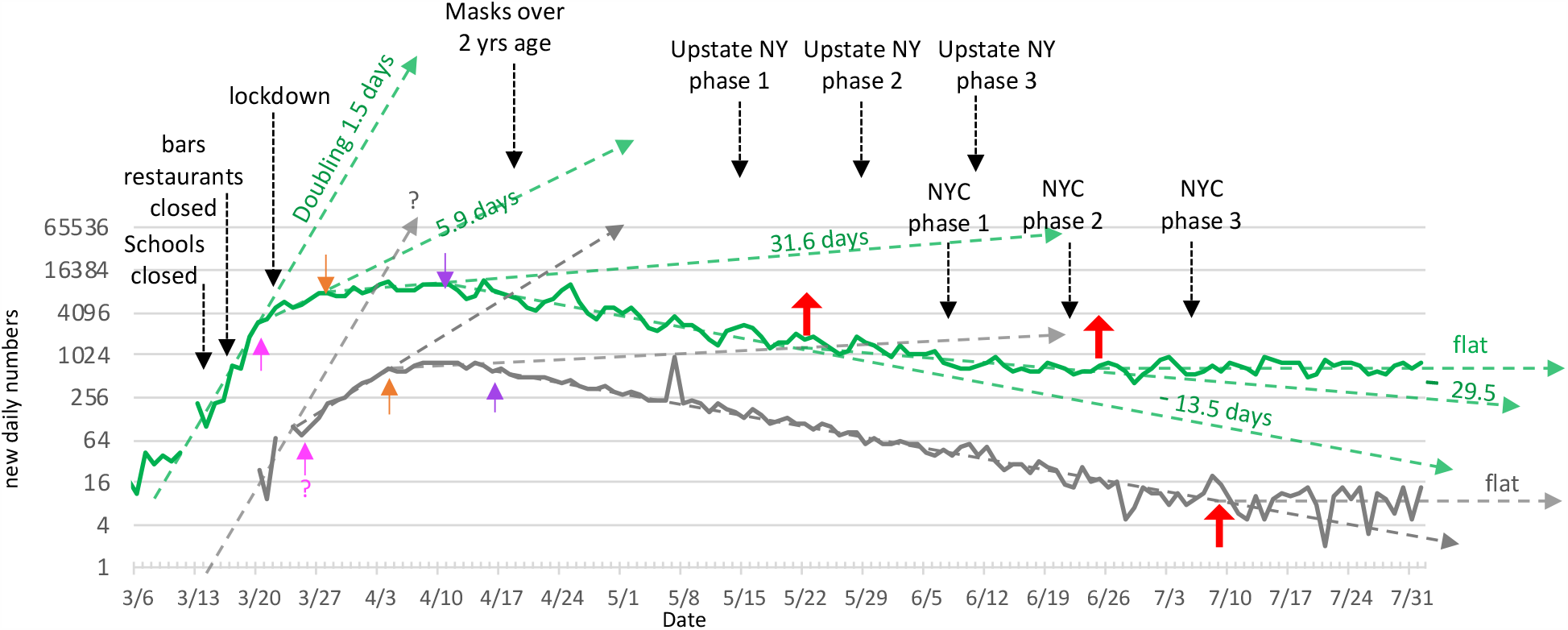
New York new daily cases and deaths: pink, orange and purple arrows point respectively to 1^st^, 2^nd^, 3^rd^ inflection points found during early stages of pandemic. Larger red arrows point to inflection points during the state’s economic openings. Question marks indicate slope and inflection point where the estimate is questionable because of lack of sufficient data points.

1. Things to note from supplemental Figure 2:
  1. Closing schools, restaurants and bars dropped the infection rate about four fold, seemingly a combination of the individual effects of closing bars and closing schools seen in states that ordered those separately.
  2. The effect of closing schools eateries is as strong here, at a time when daily new infections were close to a thousand, as they were in other states with daily new infection rates in the tens, supporting the idea that the proportional effect of a particular mandate is about the same whether implemented early or late.
  3. As with other states, opening the economy stopped the decline in infections. Opening in stages resulted in staged increases in slope.
  4. Mandating masks had no apparent immediate effect during the lockdown, as the previous mandates had already sent infection rates into decline. However, when the state opened up, it did not return to the post-school closing doubling rate of 5.9 days, as was seen in states that had not required masks (Figure 3), but instead to the lower rates that resulted from late-post-opening mask mandates. Thus, like the effect of closing schools, the effect of mandating masks seems to last as long as the mandate is in place.

## Notes

### Competing Interest Statement

The authors have declared no competing interest.

### Funding Statement

this work was entirely supported by the Division of Intermural Research of the National Institute of Allergy and Infectious Diseases, NIH

### Author Declarations

Division of Intermural Research, National Institute of Allergy and Infectious Diseases, NIH

